# Rapid Sepsis Diagnosis in Critically Ill Adults with Remoscope Label-Free Whole-Blood Imaging

**DOI:** 10.64898/2026.10.08.26364716

**Authors:** Nicholas Bratvold, Ilakkiyan Jeyakumar, Michelle Khoo, Gregory Suematsu, Olivia Chao, Tal Shwartzman, Andrew Willmore, Julia Hutchinson, Carolyn S. Calfee, Michael A. Matthay, Natasha Spottiswoode, Charles R. Langelier, Paul M. Lebel

## Abstract

Sepsis is life-threatening, yet diagnosis remains a major global challenge because existing approaches lack speed and accuracy. Here we show that label-free imaging cytometry of 10 µL whole-blood samples by Remoscope rapidly distinguishes patients with sepsis from those with non-infectious critical illness and healthy controls. In a prospective study of 29 critically ill adults, multi-physician adjudication identified sepsis (*n*=13), non-infectious critical illness (*n*=12), and indeterminate status (*n*=4); these patients were compared to healthy controls (*n*=22). A multiple instance learning (MIL) model pooled unbiased features from thousands of blood images per patient. Optimized for rule-out, the three-class model detected sepsis with 100% sensitivity and 85.3% specificity. Class activation heatmaps, unsupervised single cell embeddings, and morphometrics converged on abnormal leukocyte and erythrocyte morphology, including immature neutrophil-like cells. Label-free whole-blood imaging with MIL detects sepsis-associated hematologic signatures without stains or cold chain, establishing a foundation for rapid critical care diagnostics.

## Introduction

Sepsis is a life-threatening syndrome caused by a dysregulated host-response to infection and remains among the most urgent diagnostic challenges in acute care^1^. It causes substantial global morbidity and mortality, accounting for an estimated 20% of deaths worldwide and contributing to a large fraction of hospital deaths in the United States^2,3^. Early recognition is paramount because timely antimicrobial therapy and source control improve survival^4,5^, yet many non-infectious critical conditions often mimic sepsis including autoinflammatory diseases, endocrine crises, acute cardiovascular events, toxin exposures, or allergic reactions. At the time treatment decisions are made, clinicians must often distinguish infection-driven organ dysfunction from sterile inflammation using non-specific vital signs, routine laboratory values, and incomplete microbiologic data. This uncertainty contributes both to delayed or inappropriate treatment in patients with true infection and unnecessary antimicrobial exposure in patients without sepsis^6,7^.

Current sepsis diagnostics are poorly matched to the speed and complexity of critical care decision-making^8^. Blood cultures and other routine clinical microbiologic tests can identify causative pathogens, but they require hours to days, are often negative after antibiotic exposure, and fail to identify an etiologic pathogen in a substantial fraction of cases^9–11^. Molecular pathogen-detection methods, such as plasma metagenomic sequencing, can enable culture-independent detection, but may identify organisms of uncertain clinical relevance and have limited ability to rule out infection^12,13^. Host-response assays provide a complementary strategy by measuring the biological consequences of infection. However, conventional biomarkers such as C-reactive protein and procalcitonin have limited specificity for sepsis, while transcriptional and cellular host-response assays often require specialized sample processing, reagents, centralized testing, or analytic workflows that limit rapid point-of-care deployment^7,14–17^.

Blood cells are attractive targets for a rapid host-response diagnostic because sepsis produces systemic hematologic changes across multiple cellular compartments. Neutrophil activation, altered leukocyte morphology, immune dysregulation, platelet activation, coagulation abnormalities, erythrocyte deformability changes, and microvascular dysfunction are all features of severe infection^18–22^. Existing cellular host-response tests demonstrate that biophysical features of immune cells contain diagnostic information^15,23,24^, but most interrogate selected cell populations or require microfluidic measurements, staining, or other sample preparation. Because sepsis is a whole-blood syndrome involving leukocytes, erythrocytes, platelets, plasma factors, and flow behavior, we reasoned that rapid high-throughput imaging of native whole blood could capture a broader disease-associated cellular phenotype.

Label-free imaging cytometry provides a potential route to this goal. By imaging fresh, unstained blood directly, it preserves cellular morphology and blood-flow-associated features while avoiding stains, antibodies, lysis, centrifugation, and cold-chain dependence^25^. When coupled with machine learning, such images can be analyzed without requiring investigators to predefine which cell types or morphologic features are most informative. Multiple instance learning (MIL) is particularly well-suited to this problem because it can learn patient-level diagnostic labels from large collections of raw blood cell images, allowing disease-associated signals to emerge from heterogeneous cell populations and rare abnormal morphologies^26–28^.

Here, we evaluate the Remoscope, an ultra-rapid label-free imaging cytometry platform, as a diagnostic approach for sepsis in critically ill adults^25^. We find direct whole blood imaging signatures that accurately distinguish sepsis from non-infectious critical illness and health. Taken together, this work spotlights a new class of sepsis diagnostic that instead of direct pathogen or biomarker measurement, captures the integrated cellular consequences of infection in the systemic circulation.

## Results

### Study population

We studied 51 adults enrolled across two prospective observational cohorts and analyzed whole blood samples collected within six hours of study enrollment. The first cohort consisted of 29 critically ill adults enrolled in the Early Acute Renal and Lung Injury (EARLI) cohort who were admitted from the emergency room to the intensive care unit (ICU) with suspected sepsis, or with critical illnesses due to causes other than sepsis. The second cohort enrolled 22 healthy adults (**Figure 1a**).

**Figure 1:**
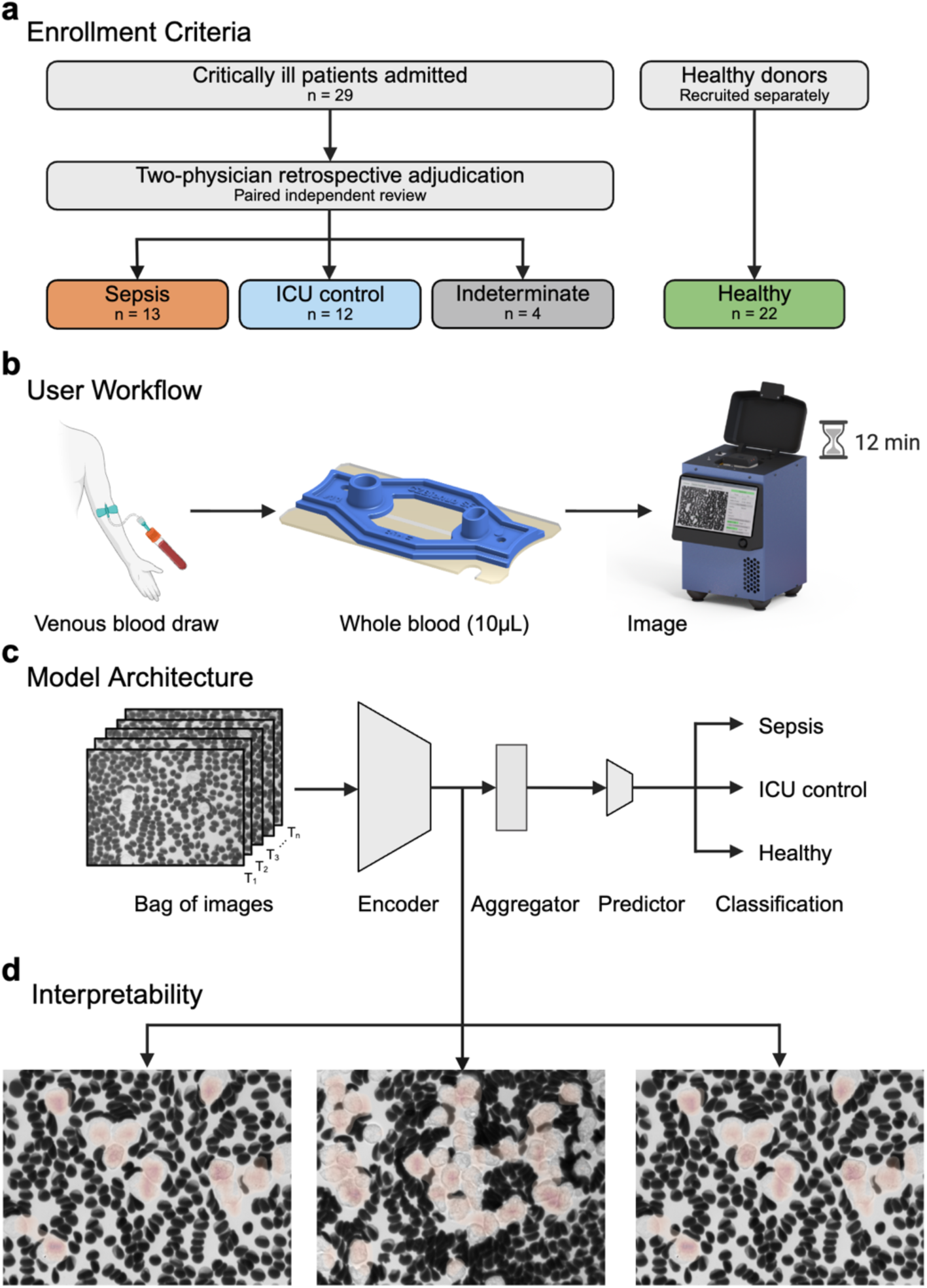
Overview of study and the Remoscope platform. ^32^. (a) Critically ill patients with acute illnesses (*n*=29) or healthy volunteers (*n*=22) were studied, and multi-physician adjudication using the Sepsis-3 criteria was employed to identify patients with sepsis (*n*=13) or non-infectious critical illnesses (*n*=12); four patients with indeterminate sepsis status were excluded. (b) 10 µL of EDTA-treated fresh whole blood was loaded into a flow cell and processed on the Remoscope without staining, lysis, dilution, or cell isolation. (c) A 3-class multiple instance learning model, termed Remo-ID, was developed for diagnostic classification of patients with sepsis, non-infectious critical illnesses or healthy controls. Remo-ID is comprised of an encoder, an aggregator, and a predictor. The encoder produces image representations, the aggregator converts a bag of encoded images into a single bag-level vector, and the predictor uses the bag vector to produce a final sample classification. (d) Example Remo-ID model attribution overlay. Red highlighted regions indicate image features influential to sepsis prediction.

Sepsis status was determined based on multi-physician adjudication of all electronic health record data following intensive care unit discharge, using the Sepsis-3 criteria and blinded to cytometry data^29^. This process identified 13 patients with sepsis, 11 of whom had a confirmed microbiologic diagnosis due to bacterial infection (8, 61.5%), viral infection (1, 7.7%), fungal infection (1, 7.7%) or bacterial-fungal co-infection (1, 7.7%) (**Table 1**). Adjudication also identified 12 patients with unequivocally non-infectious explanations for their critical illness, who served as a critically ill control population (**Supplementary Table 1**). Four individuals had an indeterminate sepsis status and were excluded from model training and performance analyses (**Supplementary Table 2**). The median age of patients with sepsis was 66.0 years (interquartile range (IQR) 46.0 – 80.0) while that of ICU controls was 55.5 years (IQR 42.8 - 64.2) and that of healthy controls 28.0 years (IQR 25.0 – 38.8). Mortality in the sepsis group was 38.5%, while that in the ICU control group was 25.0% (**Table 1).**

**Table 1:** Clinical and demographic characteristics of participants.

| Characteristic | Sepsis<br>(n=13) | ICU control<br>(n=12) | Healthy<br>(n=22) | P value<br>(ICU control<br>vs Sepsis) |
| --- | --- | --- | --- | --- |
| Patients, n | 13 | 12 | 22 |  |
| Number of experiments (support), n | 21 | 20 | 88 |  |
| Age, median (IQR) | 66.0 (46.0, 80.0) | 55.5 (42.8, 64.2) | 28.0 (25.0, 38.8) | .19 |
| Sex, Female, n (%) | 7 (53.8) | 6 (50.0) | 16 (72.7) | 1.00 |
| Race, n (%) |  |  |  |  |
| African American | 1 (7.7) | 0 (0.0) | 0 (0.0) | 1.00 |
| Asian | 3 (23.1) | 2 (16.7) | 7 (31.8) | 1.00 |
| Caucasian | 5 (38.5) | 6 (50.0) | 10 (45.5) | .70 |
| Other | 3 (23.1) | 4 (33.3) | 4 (18.2) | .67 |
| Unknown | 1 (7.7) | 0 (0.0) | 1 (4.5) | 1.00 |
| Hispanic Ethnicity, n (%) | 2 (15.4) | 2 (16.7) | 2 (9.1) | 1.00 |
| Temp Max (°C), median (IQR) | 37.4 (36.8, 38.1) | 37.2 (36.7, 37.7) |  | .51 |
| Heart Rate Max (bpm), median (IQR) | 111.0 (86.0, 126.0) | 119.0 (106.2, 125.2) |  | .62 |
| Resp Rate Max (bpm), median (IQR) | 29.0 (25.0, 32.0) | 29.5 (24.2, 33.2) |  | .83 |
| BP Systolic Min (mmHg), median (IQR) | 70.0 (64.0, 78.0) | 82.5 (69.0, 96.2) |  | .07 |
| WBC Max (10<sup>3</sup>/μL), median (IQR) | 16.3 (7.9, 28.0) | 9.4 (6.9, 13.0) |  | .07 |
| Hgb Min (g/dL), median (IQR) | 8.0 (7.3, 10.3) | 8.8 (8.1, 11.1) | 14.5 (13.7, 15.5) | .21 |
| Hct Min (%), median (IQR) | 25.0 (22.4, 31.7) | 25.9 (23.3, 32.1) |  | .46 |
| Plt Min (10<sup>3</sup>/μL), median (IQR) | 139.0 (80.2, 191.8) | 122.0 (105.5, 180.2) |  | .91 |
| Neutrophils Max (10<sup>3</sup>/μL), median (IQR) | 13.1 (5.5, 16.4) | 8.6 (6.2, 9.6) |  | .14 |
| Lymphocytes Max (10<sup>3</sup>/μL), median (IQR) | 0.6 (0.3, 1.4) | 0.6 (0.4, 1.1) |  | .79 |
| Monocytes Max (10<sup>3</sup>/μL), median (IQR) | 0.6 (0.4, 2.0) | 0.7 (0.5, 0.8) |  | .91 |
| Eosinophils Max (10<sup>3</sup>/μL), median (IQR) | 0.0 (0.0, 0.0) | 0.0 (0.0, 0.1) |  | .53 |
| Basophils Max (10<sup>3</sup>/μL), median (IQR) | 0.0 (0.0, 0.1) | 0.0 (0.0, 0.1) |  | .88 |
| Creatinine Max (mg/dL), median (IQR) | 2.1 (1.5, 2.6) | 0.9 (0.7, 1.0) |  | .03 |
| Glasgow Coma Scale, median (IQR) | 9.0 (6.0, 14.0) | 15.0 (14.2, 15.0) |  | .007 |
| APACHE III, median (IQR) | 119.0 (72.0, 131.5) | 63.5 (49.2, 74.8) |  | .01 |
| On Pressors/Inotropes, n (%) | 12 (92.3) | 6 (50.0) |  | .03 |
| Intubated, n (%) | 6 (46.2) | 3 (25.0) |  | .41 |
| Intubation Days, median (IQR) | 1.0 (0.0, 8.0) | 0.5 (0.0, 7.5) |  | .95 |
| Ventilator-Free Days, median (IQR) | 18.0 (1.0, 28.0) | 24.5 (16.8, 28.0) |  | .33 |
| Pathogen Identified, n (%) | 11 (84.6) | 0 (0.0) |  |  |
| *Bacterial* | 8 (61.5) | 0 (0.0) |  |  |
| *Viral* | 1 (7.7) | 0 (0.0) |  |  |
| *Fungal* | 1 (7.7) | 0 (0.0) |  |  |
| *Bacterial + fungal* | 1 (7.7) | 0 (0.0) |  |  |
| 28-Day Mortality, n (%) | 4 (30.8) | 3 (25.0) |  | 1.00 |
| 60-Day Mortality, n (%) | 5 (38.5) | 3 (25.0) |  | .67 |

### Rapid collection of label-free imaging cytometry data using the Remoscope

For each dataset, 10 µL of whole blood was loaded into a disposable flow cell and 20,000 images were acquired over approximately 12 minutes (**Figure 1b**). The primary dataset comprised 47 analyzed participants, 129 experiments, 2.5 million images, and 308 million unique cells (**Supplementary Table 3**).

### Evaluation of the Remoscope’s performance for sepsis diagnosis

We assessed the performance of the Remoscope platform for distinguishing patients with sepsis (*n*=13) from those with non-infectious critical illnesses (*n*=12) or from healthy controls (*n*=22). For this, we employed a three-class MIL model called Remo-ID using a ResNet-50 backbone^30^. Remo-ID treated each dataset as a bag of images and predicted a patient-level class by encoding all images in a bag, aggregating the image embeddings, then predicting on the aggregated embeddings (**Figure 1c; Methods)**. The primary analysis used leave-one-patient-out cross-validation with a fixed training epoch across folds. Model predictions across each patient’s technical replicates were averaged for evaluation. Each patient was assigned to the class with the highest score after subtracting the Youden’s *J*^31^ optimal threshold from its predicted probability.

For each clinical group, we evaluated classification performance against the two other groups combined (one-vs-rest), reporting AUC, F1 score, sensitivity, and specificity. Remoscope identified patients with sepsis with a one-vs-rest area under the receiver operating characteristic curve (AUC) of 0.98 (95% confidence interval (CI), 0.94–1.00), F1 score of 0.86 (0.67–0.97), sensitivity of 0.92 (0.75–1.00), and specificity of 0.91 (0.81–1.00). Uninfected ICU control classification achieved an AUC of 0.99 (0.95–1.00), F1 score of 0.87 (0.67–1.00), sensitivity of 0.83 (0.60–1.00), and specificity of 0.97 (0.91–1.00). Classification of healthy controls achieved an AUC of 1.00 (1.00–1.00), F1 score of 0.98 (0.91–1.00), sensitivity of 0.95 (0.84–1.00), and specificity of 1.00 (1.00–1.00) (**Table 2).**

**Table 2.** Performance metrics of MIL sepsis diagnostic classifier with 95% bootstrapped confidence intervals at Youden’s *J* threshold.

| *Class* | *Support* | *F1* | *Sensitivity* | *Specificity* | *PPV* | *NPV* | *OvR AUC* |
| --- | --- | --- | --- | --- | --- | --- | --- |
| **Sepsis** | 13 | 0.86<br>[0.67–0.97] | 0.92<br>[0.75–1.00] | 0.91<br>[0.81–1.00] | 0.80<br>[0.56–1.00] | 0.97<br>[0.90–1.00] | 0.98<br>[0.94–1.00] |
| **ICU control** | 12 | 0.87<br>[0.67–1.00] | 0.83<br>[0.60–1.00] | 0.97<br>[0.91–1.00] | 0.91<br>[0.71–1.00] | 0.94<br>[0.86–1.00] | 0.99<br>[0.95–1.00] |
| **Healthy** | 22 | 0.98<br>[0.91–1.00] | 0.95<br>[0.84–1.00] | 1.00<br>[1.00–1.00] | 1.00<br>[1.00–1.00] | 0.96<br>[0.86–1.00] | 1.00<br>[1.00–1.00] |

Of 47 participants, 43 were correctly classified with predominant errors associated with distinguishing sepsis from ICU controls. Using Uniform Manifold Approximation and Projection (UMAP)^33^, we visualized image patterns learned by Remo-ID which placed images with similar features near one another. Images grouped by clinical category with some overlap between sepsis and ICU control cases (**Figure 2c).** Remo-ID’s predictions follow these groupings (**Figure 2d)**, and coloring by sample reveals differences among patients (**Figure 2e)**. These plots suggest the model learned clinical categories despite large heterogeneity among samples from the critically ill patients. Review of per-sample errors across folds, including training data, identified the following misclassified critically ill cases: a patient with sepsis secondary to *E. coli* bacteremia in the setting of cholangitis, a polymicrobial sepsis case, a patient with ARDS secondary to bleomycin toxicity, and drug-induced colitis (**Supplementary Table 4**). Although healthy donors were younger than critically ill patients (median 28 vs 58), Remo-ID’s healthy probability was not associated with age within any group, and critically ill patients younger than 58 were all classified as non-healthy (median P(healthy) = 0.004).

**Figure 2:**
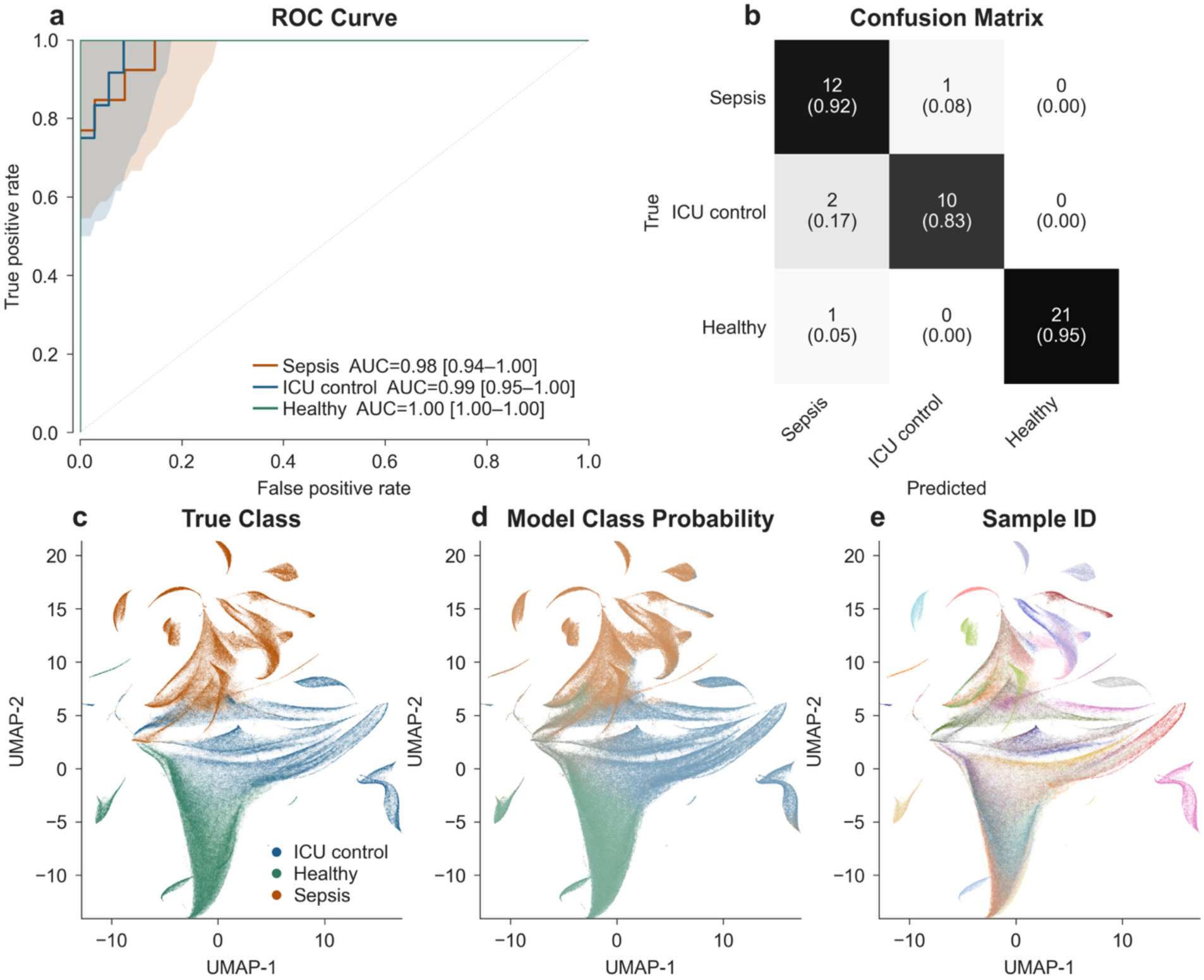
A multiple instance learning classifier accurately distinguishes sepsis, uninfected ICU controls and healthy controls. (a) One-vs-rest receiver operating characteristic (ROC) curves for the Remo-ID classifier with area under the ROC curve (AUC) and 95% confidence intervals highlighting classification performance for each group. (b) The confusion matrix shows 43 correct and 4 incorrect classifications. Image embeddings from all data collected on one Remoscope device, projected onto a Uniform Manifold Approximation Projection (UMAP)^33^ and colored by (c) true class, (d) model class probability, and (e) sample ID.

### Diagnostic performance for sepsis rule-out

The ability of a diagnostic test to accurately rule out sepsis is essential for a clinical assay seeking to guide decision making regarding the use of antimicrobials or other therapies, especially in critically ill patients. As such, we evaluated the performance characteristics of the diagnostic classifier at different decision thresholds (**Table 3**). For this analysis, a single threshold was applied to the predicted sepsis probability, whereas the class assignments in **Table 2** used per-class thresholds. At Youden’s *J*^31^, the classifier distinguished sepsis from critically ill controls with 76.9% sensitivity and 100.0% specificity; this could instead be tuned for sepsis rule-out to achieve 100.0% sensitivity at a specificity of 75.0%. For the broader sepsis versus all non-sepsis distinction, 100.0% sensitivity was achieved at 85.3% specificity, with 3/12 ICU controls and 2/22 healthy participants misclassified (**Table 3)**. Although threshold selection permitted relative prioritization of either sensitivity or specificity, F1 scores were stable over an intermediate threshold range (**Supplementary Figure 1a**; **Supplementary Figure 2**).

**Table 3.** Sepsis diagnostic performance summary at different sepsis sensitivity thresholds.

| *Classes* | *Operating Point* | *Sensitivity* | *Specificity* | *PPV* | *NPV* |
| --- | --- | --- | --- | --- | --- |
| **Sepsis vs ICU control** | Youden's $J$ | 76.9% | 100.0% | 100.0% | 80.0% |
| **Sepsis vs ICU control** | Maximum sensitivity | 100.0% | 75.0% | 81.2% | 100.0% |
| **Sepsis vs all** | Youden's $J$ | 100.0% | 85.3% | 72.2% | 100.0% |
| **Sepsis vs all** | Maximum sensitivity | 100.0% | 85.3% | 72.2% | 100.0% |

### Model interpretability and generalization

To understand the biological signals driving model predictions, we employed three complementary approaches: Gradient-weighted Class Activation Mapping (Grad-CAM)^34^ (**Figure 3**), single-cell morphological distributions using an unsupervised embedding model^35^ (**Figure 4)** and classical morphometric features derived from single-cell segmentations (**Figure 5**). Grad-CAM, which combined feature maps from the Remo-ID MIL model with target class gradients to create a visual heatmap, highlighted erythrocyte and leukocyte regions driving classification decisions (**Figure 3**). More specifically, high-confidence (p > 0.95) sepsis image regions contained a diversity of morphological cell phenotypes consistent with toxic granulation and vacuolization in neutrophils, and activated monocytes (**Figure 1**; **Figure 3c)**^36^.

**Figure 3:**
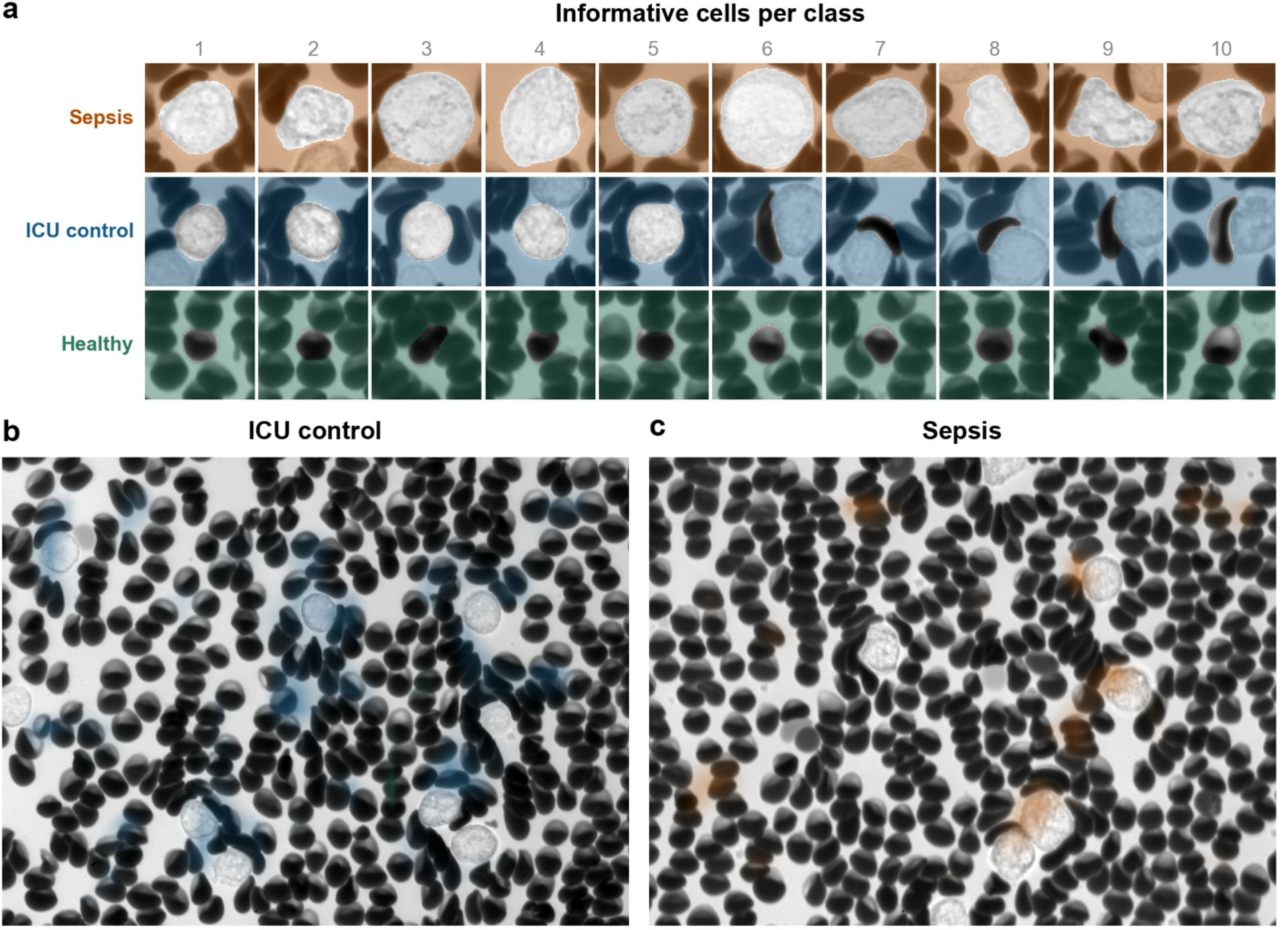
Interpretability visualization using Gradient-weighted Class Activation Mapping (Grad-CAM). (a) The most informative cells per class ranked by selecting cells near Grad-CAM predictive areas from the top 20 highest-confidence images from each experiment. In sepsis samples, highlighted cells consisted of large (3,4,6) and pleomorphic leukocytes (2,7,8,9), stress granule morphology (3,5,9,10), vacuole morphology (1,2), and band nuclei (2,7). The ICU control cells consisted of round, highly granulated leukocytes and deformed erythrocytes. The healthy samples’ highlighted cells were normal erythrocytes. (b) High-confidence (p > 0.95) ICU control image with blue Grad-CAM overlay indicating predictive areas, containing both erythrocytes and leukocytes. (c) High-confidence sepsis image with orange Grad-CAM overlay indicating predictive areas containing vacuolized and hyperpigmented leukocyte morphologies.

**Figure 4:**
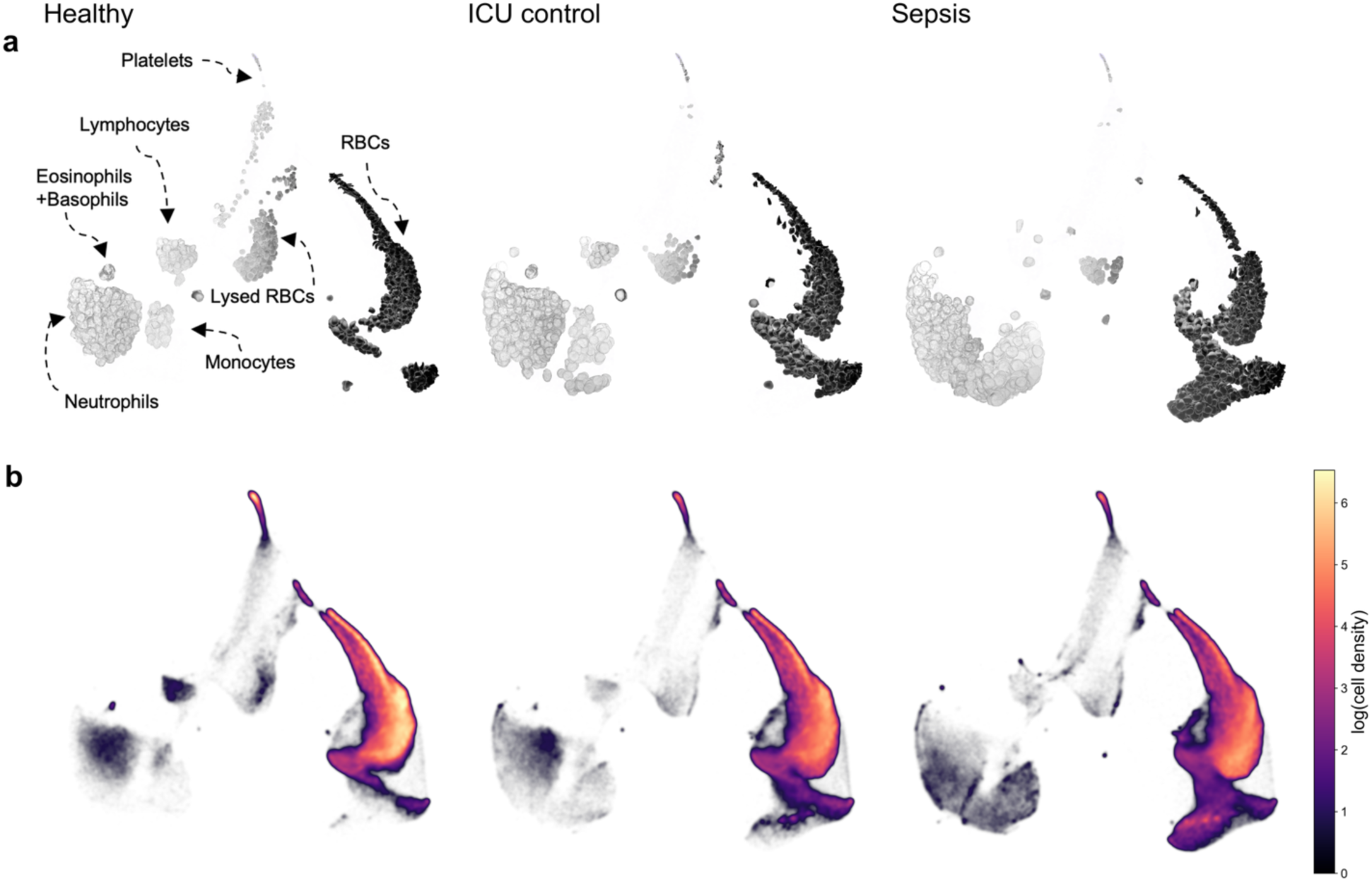
Cell morphology distribution shifts between sepsis, ICU control, and healthy groups based on unsupervised single segmented cell embeddings. ^35^. (a) Randomly sampled segmented cells, overlaid on their UMAP positions, from each of the three groups^33^. Observed morphological phenotype clusters and their proposed blood cell type correlations are shown on the healthy group panel. To class balance red vs. white cells, 2,500 large bright (area > 2300 px^2^ or mean intensity > 140/255) and 2,500 small dark cells (everything else) were gated from each group and plotted. (b) UMAPs of all cells from 100 random images from each experiment in the class, with no gating. Data points are color-coded for the logarithm of local population density.

**Figure 5:**
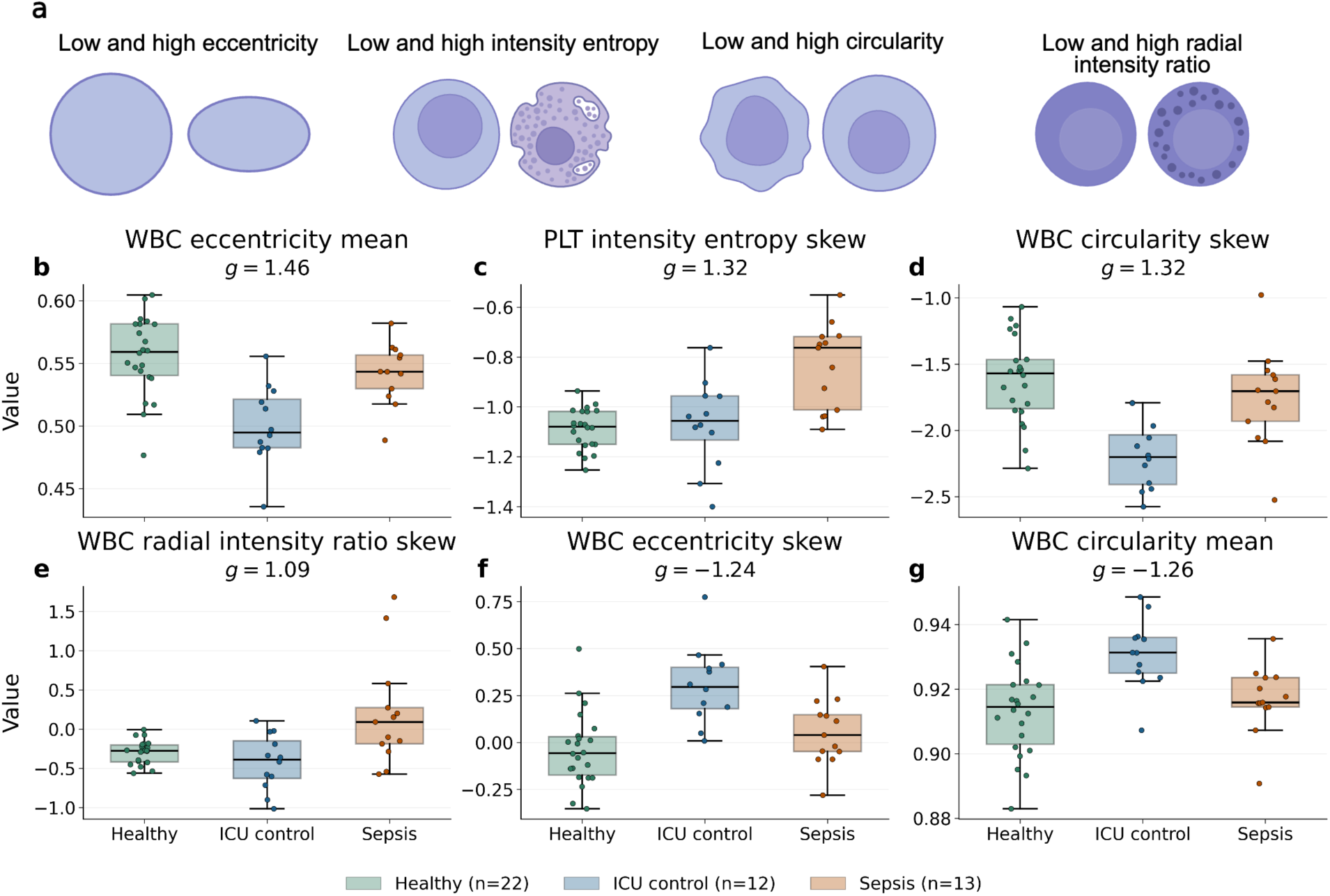
Morphometric blood cell differences between patients with sepsis, ICU controls, and healthy donors. (a) Examples of different shape and intensity parameters. (b)-(g) Titles indicate Hedges’ *g* effect size between sepsis and ICU control; displayed features have 99% confidence intervals (CI) which did not cross zero. Features are ranked by the distance from zero to the nearest CI bound (a conservative effect size estimate). Positive Hedges’ *g* denotes a higher average value in sepsis samples; negative Hedges’ *g* denotes a higher average value in the ICU control class. Distribution moments are calculated from segmented cells after partitioning into categories (RBC, WBC, platelets). Data are presented as whisker boxplots; the box extends from the first to the third quartile; center line marks the median; whiskers extend to 1.5x the interquartile range (IQR).

To further understand cellular signals associated with disease state, we analyzed population-level distribution shifts using an unsupervised single-cell embedding model^35^. Millions of segmented cells were embedded into a latent space, where population-level shifts could be compared across states of health and disease (**Figure 4**). Comparing critically ill patients to healthy controls, we noted a reduction in lymphocyte-like and an increase in neutrophil-like morphologies. Among critically ill patients, those with sepsis had a distinct population-level increase in monocyte-like cells, larger neutrophil-like cells, and immature neutrophil-like cells, denoted by thick, banded nuclei (**Figure 4**, lower left of each map). In the red blood cell (RBC) population, sepsis patients had a large increase in the fraction of macrocytic and hypochromic RBCs, consistent with previous studies reporting an increased red cell distribution width^19,20^ and hypochromic macrocytes^19^ when compared to healthy and ICU control groups.

When comparing the one influenza viral sepsis case to the bacterial sepsis cases, we observed an enrichment in cells with lymphocyte morphology alongside a decrease in cells with granulocyte morphology (**Supplementary Figure 4**), consistent with the canonical immune cell proportions associated with viral versus bacterial infections, although the complete blood count data did not demonstrate this difference, as the lymphocyte (0.76*10³/µL) and neutrophil (9.01*10³/µL) counts for the viral case were both within the interquartile range of all sepsis patients (**Table 1**).

To correlate our observations with quantitative, physiologically relevant features associated with sepsis^37–40^, we applied classical morphometric analysis to the segmented blood cell populations (**Figure 5**) and computed Hedges’ *g* effect sizes^41^ for various shape parameters across groups. The largest effect sizes between sepsis and ICU control classes were in white blood cell (WBC) eccentricity mean (*g =* 1.46; 99% CI [0.48, 3.26]; *p* = 0.0011), platelet (PLT) intensity entropy skew (*g =* 1.32; 99% CI [0.40, 2.79]; *p* = 0.0027), and WBC circularity skew (*g =* 1.32; 99% CI [0.36, 3.05]; *p* = 0.0021). Sepsis samples showed an increase in WBC eccentricity and a concomitant reduction in WBC circularity, along with an increase in skew suggesting a comprehensive shift towards a fully activated WBC population. A more positive skewness in platelet intensity entropy suggests a shift toward more activated platelets. An increase in WBC radial intensity ratio skew (*g* = 1.09) – computed as the ratio of central to peripheral pixel intensities – also suggests an elevated sub-population with higher peripheral granulation densities in sepsis as compared to both healthy and ICU control groups (**Figure 5e**).

## Discussion

Here we demonstrate that ultra-rapid, label-free imaging of unprocessed whole blood can distinguish critically ill adults with sepsis from those with non-infectious critical illness and from healthy individuals. Applying a multiple instance learning model to thousands of images per patient, the Remoscope identified sepsis with an AUC of 0.98 using leave-one-patient-out cross-validation, and when the decision threshold was tuned for rule-out, achieved 100% sensitivity at 85.3% specificity. Because the assay uses no stains, antibodies, lysis, centrifugation, or cold chain, and because classification derives from the integrated cellular consequences of infection rather than detection of a pathogen or a single analyte, it represents a point-of-care diagnostic strategy distinct from both conventional microbiologic and host-response assays.

This work demonstrates favorable performance characteristics in comparison to both existing and emerging sepsis diagnostics. For instance, automated hematology analyzers are the fastest available comparators with a turnaround time for a CBC of one minute, but lack both sensitivity and specificity^42^. Procalcitonin is among the most clinically recognized sepsis diagnostics, but has limited accuracy in critically ill populations, with a recent meta-analysis of 10 studies demonstrating a pooled sensitivity and specificity of 0.72 and 0.65, respectively^43^. Biophysical leukocyte deformability testing offers a turnaround time of 10 minutes but trades sensitivity for specificity across reported band cutoffs, ranging from 0.69/0.87 to 0.93/0.63^23^. Host gene expression assays perhaps show the most promise for achieving sepsis diagnostic AUCs > 0.90^44^, but at present require turnaround times of several hours and significant infrastructure requirements^14,44^. In comparison, Remoscope achieved an AUC of 0.98 against the combined control population, with 100% sensitivity at 85% specificity and a 12-minute acquisition, occupying a region of the speed–accuracy space that existing assays do not: near-CBC turnaround with accuracy comparable to or exceeding that of multi-hour molecular tests, and without the reagent and instrumentation dependencies those tests carry.

These diagnostic test characteristics may be particularly consequential in low-and middle-income countries (LMICs), where an estimated 85% of global sepsis cases and deaths occur^2^ and where blood culture, molecular pathogen detection, and centralized laboratory infrastructure are frequently unavailable. The Remoscope requires 10 µL of unprocessed whole blood, no stains, and no cold chain, and performs inference onboard the instrument, removing the dependencies on refrigerated consumables, reliable power for centrifugation, and network connectivity that constrain deployment of most molecular sepsis diagnostic assays. A further advantage is that the Remoscope was originally developed as a label-free malaria diagnostic, quantifying *Plasmodium falciparum* parasitemia from the same unstained 10 µL sample^25^.

Since malaria and bacterial sepsis present with overlapping febrile syndromes and treatment decisions in endemic settings are frequently made empirically without microbiologic confirmation, a single instrument reporting both parasitemia and a sepsis host-response classification could address a consequential diagnostic gap: separating patients who require antimalarials from those who require antibiotics, and identifying the subset with parasitemia who are also septic and at highest risk of death. As both classifications derive from the same image stream, this multiplexing requires no additional sample, consumable, or operator step, only an additional model.

Our data suggest that label-free image cytometry captures latent cellular morphology and flow behavior changes in whole blood that are distinct to sepsis. Grad-CAM feature maps, unsupervised cell embeddings, and classical morphometric analysis all displayed complementary patterns of leukocyte and erythrocyte morphological changes. Quantitative differences included shifts in leukocyte eccentricity, circularity, radial intensity ratio, as well as population shifts observed in unsupervised single cell embeddings (**Figure 4**). Visually, we observe several distinct morphological phenotypes, which we provisionally associate with real cell types and states, by comparing size, intensity variation, and nuclei shapes against public repositories (^36^ and **Supplementary Figure 3**). In patients with sepsis, we observed shifts toward increased endogenous cytoplasmic optical density and texture in neutrophil-like cells consistent with toxic granulation. Additional putative morphological associations include: increased rouleaux formation, leukocyte vacuolization, increased area distribution width of erythrocytes and leukocytes, increased numbers of monocytes, reticulocytes, erythroblasts, hypochromic erythrocytes, macrocytes, hemolytic inclusions, and band neutrophils. Because cells experience moderate shear and planar confinement on the Remoscope, morphological features observed in these data may not be directly comparable to other methods; it is also not yet understood to what extent cellular rigidity influences shear distortion rates. Future quantification of these deformation patterns could clarify their relationship to blood rheology and their contribution to classification. Collectively, our data suggest that sepsis is characterized by heterogeneous changes throughout whole blood, and that the novel utility of this method lies in its ability to capture a breadth of features across blood compartments.

Visualization of the embedded image feature maps (**Figure 2c-e**) demonstrated substantial heterogeneity across critically ill patients in our study, suggesting the data may contain further condition-specific information. Given that the Remoscope device images whole blood in moving liquid form within an ultrathin flow layer, it may capture population-level flow field features that are not accessible in conventional smear workflows or single-cell deformability measurements. In addition to sepsis and malaria, the Remoscope platform could also enable detection of red blood cell dyscrasias (e.g., sickle cell disease, thalassemia, hereditary spherocytosis), leukemias, or certain inflammatory conditions with distinct blood cell morphological perturbations (e.g., hemophagocytic lymphohistiocytosis). With millions of cells imaged from each sample, it is also possible to detect rare cells or weak signals that would be undetectable in lower throughput assays. These findings motivate follow-up studies linking model outputs to blinded hematopathology reviews, transcriptional profiling, and clinical laboratory test values.

This study has several strengths including development of a novel, pathogen-agnostic method for rapid sepsis diagnosis, deep clinical phenotyping including rigorous adjudication of sepsis, a prospective study design, and a platform that is low cost and conceivably deployable in clinical settings worldwide. This study also has limitations including a limited sample size, a single-site design, lack of an independent external validation cohort, a young healthy control group, and limited representation of viral sepsis. Our cross-validation approach prevented data leakage during model training, but may not have eliminated all instrument effects, sampling errors, and outlier vulnerability.

Independent validation of these findings will be necessary to assess the generalizability of results. Future work connecting Remoscope data with immunoprofiling and microbiologic data could advance our understanding of sepsis biology. Finally, future studies in pediatric and neonatal populations, as well as in malaria-endemic regions where febrile illness diagnosis is most consequential and syndromic overlap is common, would significantly extend the global utility of label-free imaging for rapid infectious disease diagnosis.

## Methods

### Study design

We studied adults admitted from the emergency room (ER) to the ICU who were prospectively enrolled in the EARLI observational cohort at the UCSF Moffitt-Long Hospital between September 2025 and May 2026. Samples were collected within 24 hours of ICU admission or in the ER. Exclusion criteria for this study included hospice or comfort care patients, patients exclusively with trauma, neurological or neurosurgical reasons for ICU admission, membership in a group considered vulnerable (pregnancy, minor status, or prisoner status) or an anticipated ICU stay <24 hours. Sepsis adjudication was performed following ICU discharge or death according to the Sepsis-3 criteria and carried out by two physicians with full access to all electronic medical record data. Samples from patients with indeterminate sepsis status were excluded from model training and performance analyses. Healthy donors were pre-screened using an eligibility questionnaire to ensure the absence of active infections, hematological abnormalities such as malignancies, or travel to malaria-endemic regions.

### Ethics statement

The IRB of the University of California, San Francisco gave ethical approval for this work under protocol 10-02852, which granted a waiver of initial consent for blood sampling of critically ill adult patient in the Early Assessment of Renal and Lung Injury (EARLI) cohort, after which informed consent was obtained from patients or surrogates^45,46^. The University of California, San Francisco gave ethical approval for the analysis of blood from healthy adult donors under IRB protocol 22-36117.

### Sample processing and Remoscope device operation

Whole blood samples were drawn by venipuncture from EARLI study participants or healthy controls into EDTA-coated collection tubes (Greiner Bio-One 22040161) or (BD # 02-687-98), respectively. Remoscope flow cells were primed with 1% w/v Poloxamer-188 solution in Tris-Borate-EDTA buffer to enhance surface wetting, then loaded with 10 µL of whole blood within six hours of collection. Each acquisition run consisted of 20,000 images collected over approximately 12 minutes, comprising approximately two million unique cells. Each cell was imaged an average of four times, for a total of approximately eight million cell detections per run. In the EARLI cohort, if more than 20 µL of blood was available, a second technical replicate was acquired. Due to the availability of larger volumes of healthy donor blood, a total of four technical replicates were acquired for the healthy control samples.

### Multiple Instance Learning (MIL) model

We implemented a form of weakly supervised classification using a MIL framework we termed Remo-ID. In the dataset, labels (healthy, sepsis, ICU control) were available at the patient level. MIL does not require a label at every image or cell; instead, it assigns a single label to a bag, which is a collection of images drawn from the same individual. The model learns to predict the patient label from its bag of images, allowing unbiased and data-driven disease state feature discovery.

#### MIL architecture

Remo-ID treated each Remoscope experiment as a bag of images and produced an experiment-level prediction. While a single Remoscope experiment consisted of 20,000 raw images, due to compute constraints, subsets were sampled during training to create the bags.

The model encoder consisted of a ResNet-50 backbone pretrained on the ImageNet dataset. ResNet-50 is a commonly used convolutional neural network (CNN) architecture that has pretrained weights for general RGB image classification^30,47^. The pretrained model’s broad exposure to a diversity of image features was leveraged as a starting point to reduce compute burden during training. The model was adapted for single-channel greyscale input by replacing the first convolutional layer. Each image in a bag was encoded into a feature embedding, and the embeddings were aggregated into a single bag-level representation using a gated attention-pooling mechanism^28^. A classifier head mapped the pooled representation to the bag-level class (**Figure 1c)**. During training, the final two ResNet-50 blocks and classifier head were fine-tuned.

#### MIL training procedure

Each bag contained 50 images at 1032 x 772 pixels. Twelve bags, four of each class, were accumulated per weight update, creating an effective batch size of 600 images. Images were augmented with randomized horizontal flipping and masking to eliminate any spurious lateral bias in cell distributions and to ignore stuck cells and debris. When expanding across multiple instruments, additional augmentations may be included to reduce machine to machine differences (**Supplementary Note 1).** Training was limited to a maximum of 50 epochs of 100 batches: 60,000 images/epoch, three million images and approximately one billion cells per full training run. Parameters were optimized with AdamW^48^, and loss was computed with categorical cross-entropy. Training took place on a single GPU (any of A40, A100, H100, or H200) and 14 CPU cores. On average, a single fold used 2 GPU hours and 40 CPU minutes.

### MIL inference

At inference, 500 images were randomly sampled from an experiment to form a bag. The larger bag size reduced per-experiment sampling variability (**Supplementary Figure 5**). The bag was passed through Remo-ID to obtain experiment-level class probabilities, from which the most likely class was selected. Predictions were generated for five independently sampled bags and averaged to obtain a final experiment-level prediction. For the primary analysis, probabilities from each participant’s technical replicates were averaged before patient-level evaluation.

### MIL cross-validation approach

To safeguard against performance overestimation and overfitting on a small dataset, we used a leave-one-patient-out cross-validation strategy, ensuring all replicates from the same patient/donor were always assigned to the same fold. We selected leave-one-patient-out to maximize the amount of training data and eliminate the risk of evaluating on a lucky train-test split by ensuring every sample is held out once^49^. All 47 folds were trained for the same fixed number of epochs to avoid biased stoppage conditions^50^. Performance metrics and class thresholds were computed on the pooled held-out predictions with 95% confidence intervals estimated by bootstrap resampling (1,000 repeats). Unless otherwise stated, class thresholds were selected with Youden’s *J* threshold optimization^31^.

### Classical morphometric analysis

Segmentation masks, which separate pixels belonging to cells from the background, were generated for each experiment using fine-tuned Cellpose-SAM^51^. Each experiment consists of 9M segmented objects on average (**Supplementary Table 5**). For each segmented object, region properties (area, perimeter, eccentricity, solidity, and pixel intensity statistics) were computed using scikit-image^52^. We additionally computed the intensity entropy, which is the Shannon entropy of the pixel values of an object. We define the radial intensity ratio as the ratio of the mean pixel values at or less than the median radius of an object to those in the outer half, which serves as a proxy for peripheral granulation and intensity homogeneity. Objects were first partitioned into RBCs, WBCs, and platelets by area-intensity criteria. An additional eccentricity maximum filter was applied to all objects smaller than 400 px^2^ to remove segmentation artifacts.

Each category’s region properties were summarized using standard distribution moments (mean, standard deviation, and skew), forming a unique morphological feature matrix for each sample. We computed pairwise Hedges’ *g* effect sizes^41^ between sample classes to identify differences across multiple feature dimensions, and 99% confidence intervals (CI) via bootstrapping with *n*=20,000. We highlight six features with the largest distance between their nearest confidence interval and zero and whose confidence intervals do not cross zero (**Figure 5)**. *P* values are reported using an exact permutation of Welch’s *t*-statistic over the entire null hypothesis label distribution as the fraction of assignments yielding a magnitude at least as large as the observed^53^.

## Data and code availability

All data and analysis code are being prepared for open-source publication.

## Data Availability

All data and analysis code are being prepared for open-source publication.

## Acknowledgments

The authors would like to acknowledge UCSF and Biohub for laboratory space, supplies, and funding. We thank Genay Pilarowski, Cristina Tato, Kelsey Zorn, and Sukhman Sidhu for assisting with the healthy donor blood collection. We are very grateful to Rafael Gomez-Sjoberg and Joe DeRisi for their support and mentorship.

## Supplementary Material

### Supplementary Note 1: Generalization across instruments

Images acquired by the Remoscope are highly consistent due to autofocus, auto-brightness, and auto-flow adjustment, ensuring variation within a single device is dominated by the biological content of the blood sample. Due to the sheer number of images and cells in comparison to the number of participants, strong data augmentation is unnecessary as there is enough meaningful natural variation in the images. However, we observe scope-to-scope batch effects caused by variation in objective quality, illumination patterns, focus plane tilt, and dust particles on the camera sensor. When expanding across multiple Remoscope devices, additional augmentations may be used to replicate scope batch effects such as:

● **Additive random noise** to prevent image memorization.
● **Gradient Gaussian blur** along a random 3D vector to simulate focal plane variation and slight camera-to-sample tilt.
● **Intensity jitter** to account for small brightness fluctuations while preserving hematocrit signals in red blood cells.
● **Background subtraction and swapping** via an experiment 99% maximum intensity projection to remove scope-specific illumination patterns and stuck debris.
● **Random masking of image and latent space** to reduce debris memorization and drive a richer and more robust representation.

No spatial rescaling was applied which is common in image augmentation pipelines as absolute cell size is a diagnostically relevant feature.

## Supplementary Figures

**Supplementary Figure 1:**
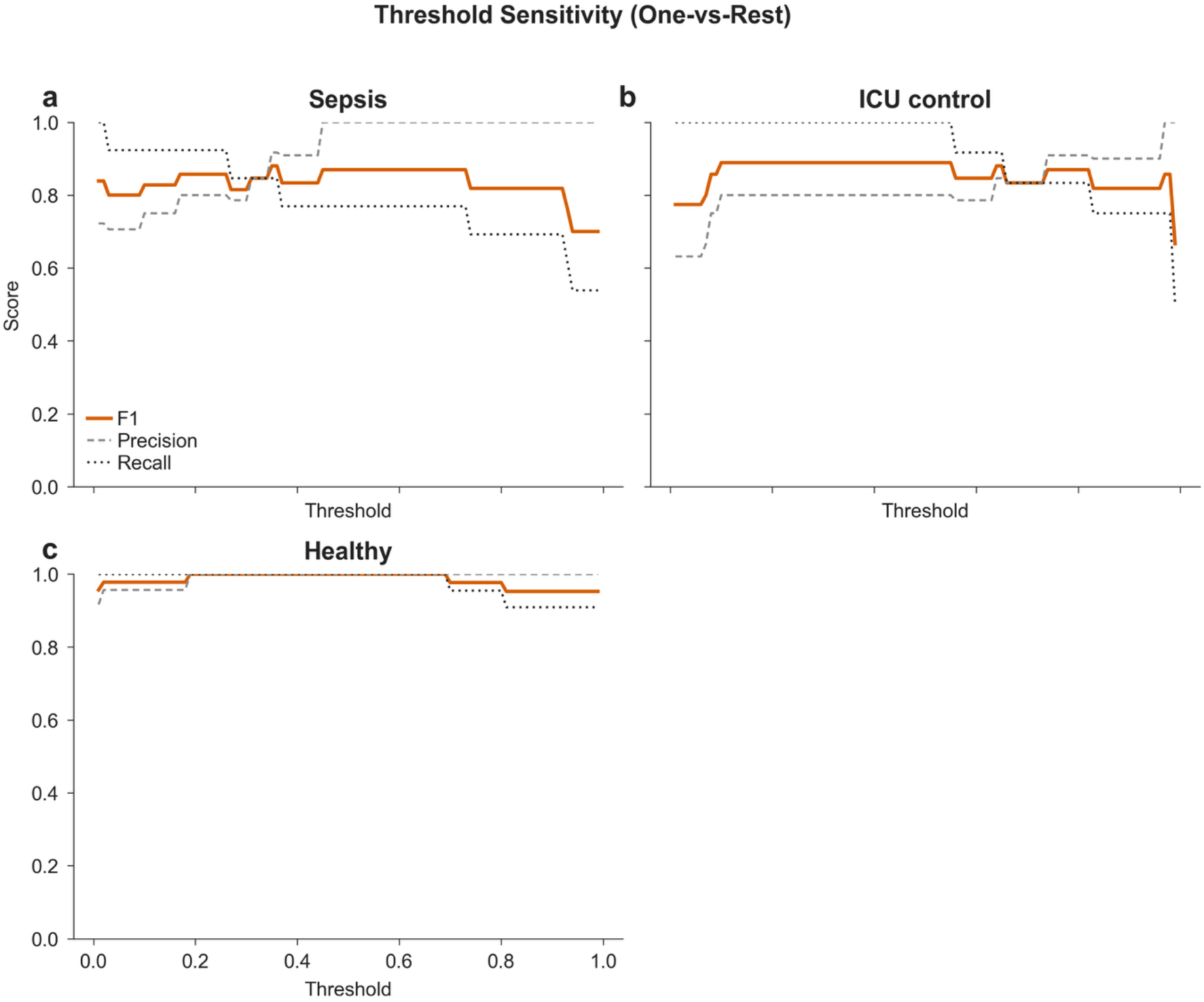
Threshold sweep of one versus rest F1, precision, and recall characteristics of sepsis, ICU control, and healthy classes.

**Supplementary Figure 2:**
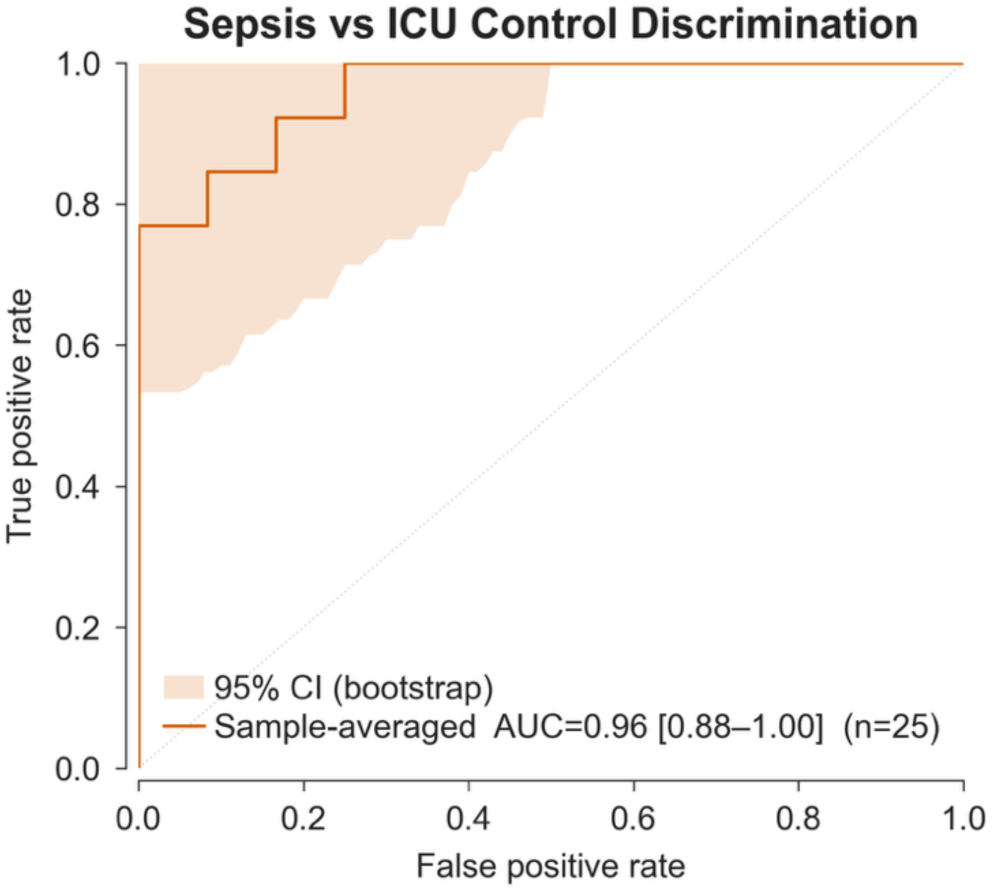
Binary sepsis versus ICU control ROC curve. The orange curve with 95% bootstrap confidence intervals (CI) demonstrates an AUC of 0.96 [0.88-1.00] in 25 critically ill patients.

**Supplementary Figure 3:**
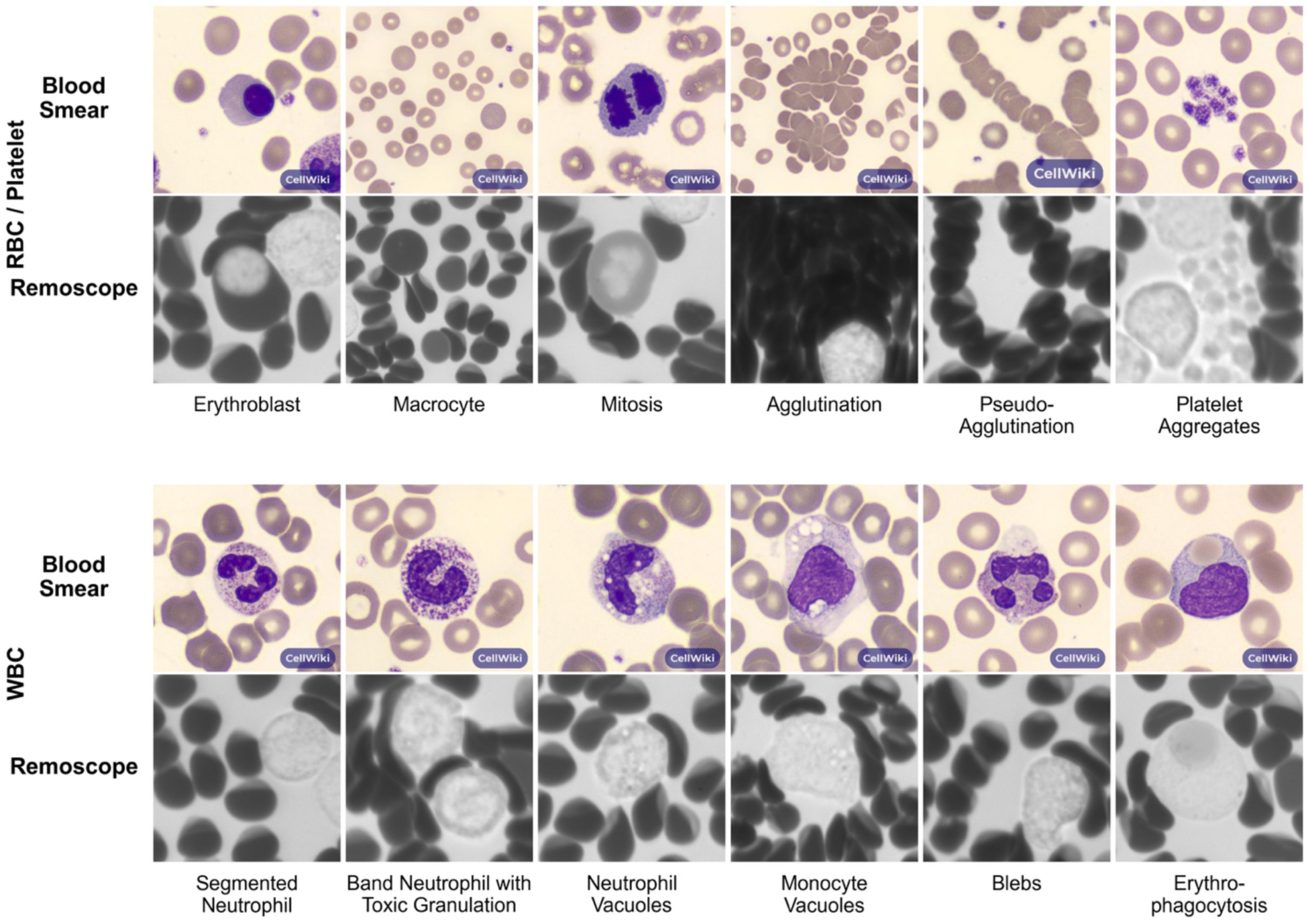
A hand-curated, putative matchup of publicly available Giemsa-stained thin blood smear examples. ^36^ **with Remoscope images from sepsis patients.** The top row contains abnormalities related to red blood cells and platelets. The bottom row contains various abnormal white blood cells. We hypothesize that the pigmented granules appearing darker in the label-free Remoscope images correspond to toxic granulation seen in stained images.

**Supplementary Figure 4:**
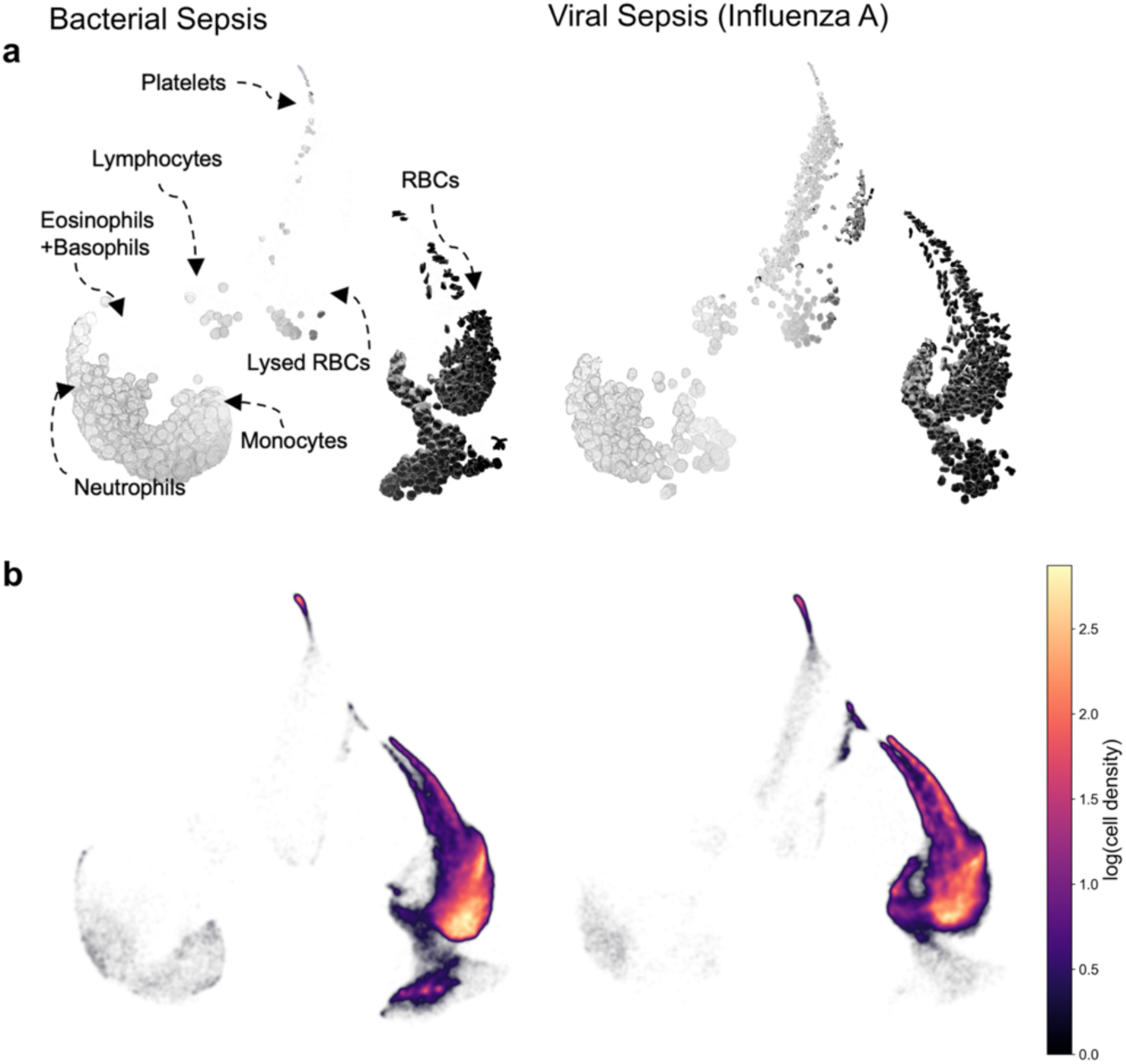
UMAP projections of unsupervised single segmented cell embeddings comparing bacterial versus viral sepsis cases. (a) Randomly sampled individual segmented cells, arranged on their UMAP positions, from a pool of Gram-positive bacterial sepsis cases and one viral (Influenza A) sepsis case. To class balance red vs. white cells, 2,500 large bright and 2,500 small dark cells were gated from each class and plotted. (b) Conventional UMAPs of all cells from 100 random images from each experiment in the class, with no gating. The bacterial density UMAP was subsampled down to 375,000 points to match the number of points in the viral density UMAP. Data points are color-coded for the logarithm of local population density.

**Supplementary Figure 5:**
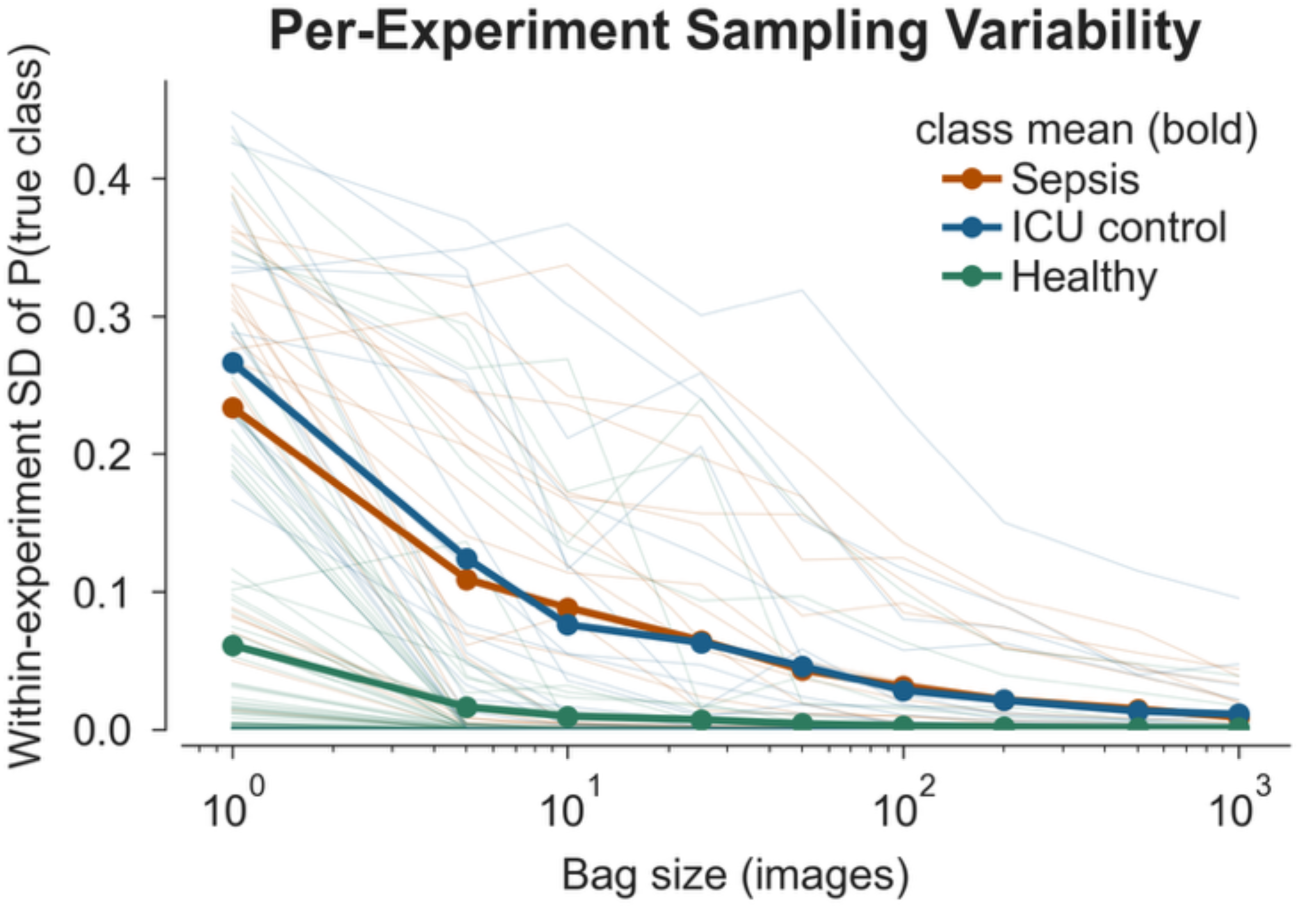
Per experiment sampling variability as a function of bag size plotted on a linear-log plot. Thin lines show variability for individual experiments and bold lines are per-class means. At each bag size, 20 bags are randomly sampled and evaluated on, and a standard deviation is computed.

## Supplementary Tables

**Supplementary Table 1:** Clinical diagnoses of ICU control group. 1 (8.3)

| **Clinical Diagnosis** | **N (%)** |
| --- | --- |
| ARDS from bleomycin toxicity | 1 (8.3) |
| Acute heart failure and SLE nephritis | 1 (8.3) |
| Drug-induced colitis | 1 (8.3) |
| HLH flare and T cell lymphoma | 1 (8.3) |
| Hypertensive emergency and emesis | 1 (8.3) |
| Hypoxemic respiratory failure due to acute cellular rejection of lung transplant | 1 (8.3) |
| Interstitial lung disease, post lung transplant | 1 (8.3) |
| Large volume hematemesis | 1 (8.3) |
| Hypoxemic respiratory failure due to MDA5 interstitial lung disease | 1 (8.3) |
| Metastatic oropharyngeal squamous cell carcinoma | 1 (8.3) |
| Metastatic hemangiopericytoma | 1 (8.3) |
| SLE-related non-infectious endocarditis | 1 (8.3) |

**Supplementary Table 2:** Indeterminate sample model predictions. The four indeterminate samples were evaluated by every model in each fold.

| **Indeterminate Samples** | **Clinical Diagnosis** | **Microbiology** | **Predicted Class** | **Fold Agreement (47 folds)** |
| --- | --- | --- | --- | --- |
| **2331** | Meningoencephalitis following respiratory infection weeks prior. | M. pneumoniae serology | ICU control | 89% |
| **2332** | EtOH withdrawal, acute pancreatitis, cirrhosis complicated by spontaneous bacterial peritonitis. | — | ICU control | 100% |
| **2338** | Acute on chronic hypercarbic respiratory failure, heart failure with preserved ejection fraction, pulmonary hypertension. | C. freundii from urine culture. | Sepsis | 100% |
| **2342** | Lung transplant hospitalized for hypoxemic respiratory failure thought to be 2/2 pneumonia versus chronic lung allograft dysfunction. | Aspergillus | ICU control | 94% |

**Supplementary Table 3:** Dataset demographic and support summary.

| **Class** | **Patients** | **Experiments** | **Images** | **Cells** | **Approximate deduplicated cells** |
| --- | --- | --- | --- | --- | --- |
| **Sepsis** | 13 | 21 | 420,000 | 175,686,370 | 43,921,593 |
| **ICU control** | 12 | 20 | 387,475 | 179,932,463 | 44,983,116 |
| **Healthy** | 22 | 88 | 1,756,987 | 876,733,439 | 219,183,360 |
| **Total** | 47 | 129 | 2,564,462 | 1,232,352,272 | 308,088,068 |

**Supplementary Table 4:** Summary of Remo-ID model errors by patient sample and physician diagnosis.

| **Experiment** | **Participant** | **Error type** | **Clinical diagnosis** | **Pathogen** |
| --- | --- | --- | --- | --- |
| **20250918135444** | 2308 | Sepsis → ICU control | Polymicrobial sepsis | *K. pneumoniae*,<br>*M. morganii*, *E. faecalis*, *C. amalonaticus*,<br>*P. mirabilis* |
| **20260130144403** | 2328 | ICU control → Sepsis | ARDS 2/2 bleomycin toxicity | - |
| **20260209155459** | 2330 | Sepsis → ICU control | Sepsis secondary to *E. coli* bacteremia in the setting of cholangitis | *E. coli* |
| **20260407111233** | HBDB-140 | Healthy → Sepsis |  | - |
| **20260407110915** | HBDB-140 | Healthy → Sepsis |  | - |
| **20260407135412** | HBDB-149 | Healthy → Sepsis |  | - |
| **20251218125729** | 2320 | ICU control → Sepsis | Drug-induced colitis | - |

**Supplementary Table 5:** Segmented object counts of experiments used in morphometric analysis.

| *Class* | *Number of runs* | *Median object count* | *Minimum* | *Maximum* |
| --- | --- | --- | --- | --- |
| *Healthy* | 22 | 10,178,802 | 8,943,995 | 11,753,237 |
| *ICU control* | 20 | 9,022,422 | 3,568,817 | 11,434,195 |
| *Sepsis* | 21 | 8,342,274 | 6,231,629 | 11,978,955 |

